# Field Observations on the Use of Rapid Diagnostic Tests for Human African Trypanosomiasis in Nigeria

**DOI:** 10.1101/2025.06.21.25330043

**Authors:** Chinwe U. Chukwudi, Omolara S. Adenekan, Jonadab C. Chinwendu, Amadea O. Enwonwu, Chinonso C. Iyi, Uchenna H. Anyaorah, Chukwuebuka V. Ugwu, Michael O. John, Glory N. Joseph, Akwoba J. Ogugua, Silva M. Anika

## Abstract

**Introduction:** Human African Trypanosomiasis (HAT) is a Neglected Tropical Disease targeted for elimination by 2030. Control efforts have reduced prevalence, but diagnostic challenges for field surveillance persist. Rapid Diagnostic Test kits (RDTs), used as frontline diagnostic tools for field surveillance, are often based on Variant Surface Glycoproteins (VSGs), which undergo constant mutations and exhibit considerable geographical diversity. Some VSGs are absent in Cameroon and Nigerian trypanosome strains/isolates.

**Method:** This study evaluated the reliability of HAT RDTs using human and animal blood samples from Nigeria. Seropositive samples from ELISA and CATT were tested with Abbott Bioline HAT 2.0 RDT.

**Result:** All human samples tested negative, while some animal samples were positive on the RDT. This raises concerns about the reliability/suitability of this RDT for HAT field surveillance in Nigeria.

**Conclusion:** This study emphasizes the importance of incorporating diverse Trypanosoma strains into RDT development and ensuring validation across all endemic areas for effective field surveillance and disease control.

## Introduction

Human African Trypanosomiasis (HAT), also known as sleeping sickness, is caused by two subspecies of *Trypanosoma brucei: Trypanosoma brucei gambiense* (*Tbg*), which causes gHAT in West and Central Africa, and *Trypanosoma brucei rhodesiense* (*Tbr*), which causes rHAT in East and Southern Africa. The parasite is transmitted by tsetse flies of the *Glossina* species (1). gHAT is characterized by a long incubation period (up to several years), chronic/intermittent fever and low parasitemia in the early stage; and severe neurological symptoms in the advanced stages. Some infected people remain asymptomatic (2). These features complicate early diagnosis and treatment (3). HAT primarily affects vulnerable populations in remote and underserved areas with limited access to healthcare infrastructure (4). Despite successful control efforts leading to reduced prevalence, disease detection is still challenging due to inadequate diagnostic tools, especially in rural regions where advanced diagnostic techniques are inaccessible.

Field surveillance of HAT is hindered by limited availability of field diagnostic tools, and low sensitivity/specificity of available serologic assays. These challenges can result in false negatives and a false sense of low or no disease prevalence, further truncating control efforts. Rapid Diagnostic Tests (RDTs) have been integrated into surveillance programs to enhance disease detection (5). Its sensitivity and specificity, which are crucial for accurate diagnosis, vary across regions due to differences in parasite strains and geographic locations (6,7). While these RDT kits offer practical solutions for field use, concerns persist about their reliability, especially in some low-prevalence settings with atypical isolates and Variant Surface Glycoprotein (VSG) patterns.

The Card Agglutination Test for Trypanosomiasis (CATT) is a widely used tool for field screening due to its simplicity and quick turnaround (8). It detects antibodies against the variant surface glycoprotein (VSG) Litat 1.3. However, this antigen is absent in some *T. b. gambiense* strains, including those found in countries like Cameroon and Nigeria (9,10). This limits the test’s effective use in disease monitoring due to concerns for false negatives in these areas.

The Abbott Bioline HAT 2.0 RDT was developed as an improvement over earlier versions and detects antibodies using two recombinant antigens: invariant surface glycoprotein (ISG) 65 and VSG LiTat 1.5 (5). While it offers advantages such as affordability and field applicability, sensitivity and specificity remain an issue, particularly in malaria-endemic regions like Nigeria (5–7).

## Method

Blood samples were collected in EDTA-coated sample bottles from hospital diagnostic laboratories (for human samples) and abattoirs (for animal samples) from two states in Southern Nigeria (Delta and Enugu States). The samples were transported to the lab on ice packs, and serum was extracted by centrifugation. The serum samples were first screened for HAT with ELISA using plates coated with native antigens. The samples that tested positive on ELISA at 1:40 and 1:200 serum dilution were then screened with CATT kit (obtained from the Institute of Tropical Medicine, Belgium) at 1:4, 1:8, 1:16, 1:32, 1:64 serum dilutions. Then, those samples that tested positive on both ELISA (1:200 dilution) and CATT (1:64 dilution) were further tested with an RDT kit (Abbott Bioline HAT 2.0), which was obtained from the Federal Ministry of Health.

## Results

A strong agglutination was observed at various serum dilutions for the ELISA-positive human and animal samples when tested with CATT (Figure 1). However, when the CATT-positive samples were tested on the RDT kits, none of the human samples were positive but two of the animal samples (both from pigs) were positive on the RDT kit (Figure 2). A summary of the samples tested and the results from all serologic tests done is shown in Table 1.

**Table 1.**
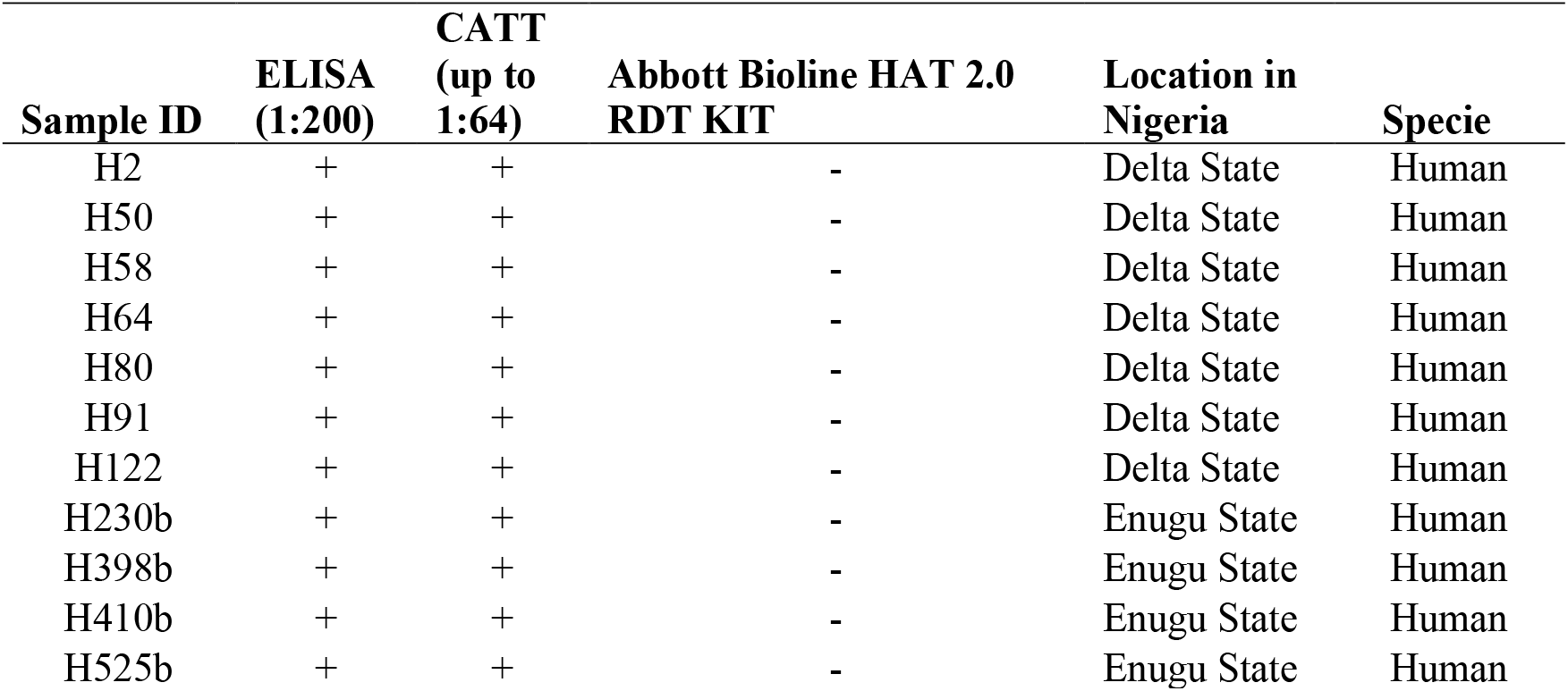
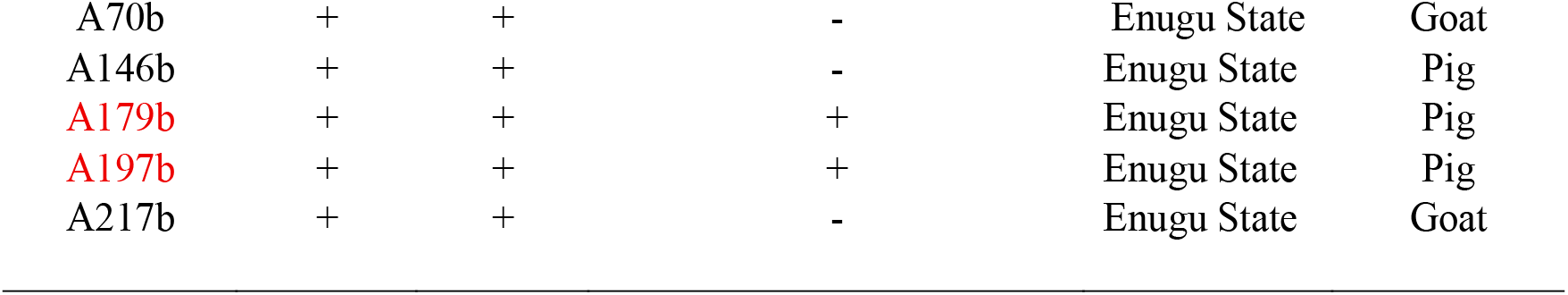
Summary of test results for human and animal samples using different serologic assays. All human and animal samples tested negative on Abbott Bioline HAT 2.0 RDT KIT except for two animal samples, A179b and A197b.

**Figure 1.** Representative results of CATT for Human and Animal samples following positive ELISA. Samples showed positive up to 1:16. **A**lt text: Images of CATT assays of ELISA-positive serum samples showing agglutination (positive
results) in all human and animal samples.

**Figure 2:** Representative results of Abbott HAT 2.0 RDT Kit for human and animal samples that were positive on ELISA and CATT assays. Arrow indicates positive result at test line **1**. **A**lt text: Images of HAT RDT kit assays of seropositive human and animal samples showing positive result on test line 1 for only 2 of the animal samples and no positive human sample.

## Discussion

Nigeria is considered a HAT endemic country. However, no cases have been reported in the WHO database from Nigeria since 2012, except one case that was diagnosed in 2016 from a Nigerian visiting the United Kingdom (3). It is highly unlikely that this UK-diagnosed case is an isolated infection, hence raising questions about HAT diagnosis and reporting in Nigeria. There is very little HAT surveillance or epidemiological data available from Nigeria. This lack of HAT surveillance data from Nigeria is a complex combination of several factors, including the lack of appropriate diagnostic tools, inefficient disease reporting protocols, and little/no surveillance activities by health authorities. While indigenous researchers report case detection using several serologic, molecular, and biological means (2,12)(13), these reports are hardly validated by the health authorities for appropriate disease reporting. WHO largely relies on a serologic test (trypanolysis) for the validation of detected cases before official acceptance for reporting despite the drawbacks of these serologic tests (10,14). It is interesting to note that even the WHO-recorded case of 2016 was also negative in a trypanolysis test (3). It has been demonstrated that *Trypanosoma brucei gambiense* VSGs and immune-reactivity tests are clearly correlated with geographical origin, and Nigerian samples have exhibited variable responses to immunologic/serologic tests as the targeted Variable Antigen Type (VAT) for these tests are known to be absent in isolates from certain geographical locations such as Nigeria and Cameroon (9,10). It is becoming apparent that variations in local strains, as indicated by the genetic diversity of *Trypanosoma* species across different geographical regions, may compromise the effectiveness of serologic tests generally, including RDTs (9). These serologic tools, which were developed and validated elsewhere, may not reliably detect Nigeria’s local parasite strains, thereby increasing the risk of false negatives (5,6). This RDT kit has previously been shown to perform poorly with unreliable results in passive surveillance studies in Nigeria (15). However, it is still being used for passive case detection by the health authorities. This could contribute largely to the inability to detect cases in Nigeria, leading to misleading prevalence reports/data for policy implementation (16). Hence, there is a need to take this geographic diversity of *Trypanosoma* strains into consideration in the development of diagnostic tools.

The usual concern for HAT RDTs in many endemic areas is the occurrence of false positive reactions, which are believed to be associated with cross-reacting antibodies in malaria co-endemic areas.(5,17). In high-prevalence settings, a test with low specificity will produce more false positives. Hence, increasing specificity ensures that positive results are more likely to be true cases. This improves the positive predictive value (PPV) which is the likelihood that a person who tests positive actually has the disease. It is widely believed that RDTs using recombinant antigens have higher specificity than those with native antigens (6,18). However, the findings in this report show that this may compromise sensitivity in some cases, especially in low-prevalence settings where a higher sensitivity is more desirable to ensure that all positive cases are detected. This is particularly important when disease elimination is targeted, as is the case for HAT. Nigeria has low HAT prevalence, and the use of a low-sensitive surveillance tool like the RDT kit portends great dangers for false negatives, thereby increasing the risk of missed infections, continued disease transmission, and the possibility of disease/epidemic re-emergence.

Although these serologic tests have been acclaimed to be specific for the detection of human infection by *T. brucei gambiense*, Ilboudo et al had demonstrated using pigs that the VATs used in the trypanolysis test is not specific for Tbg infection (14). The positive test results in animal samples obtained in this study raises important questions about the specificity of these diagnostic tools. One may argue that it is possible these positive animal samples were infected with Tbg. But the question remains as to why the RDT kit is not detecting the human samples that were also positive on ELISA and CATT like the animal samples. On the other hand, the animal samples were positive in the invariant surface glycoprotein (ISG) test line of the RDT (test line 1), and it is possible that this antigen is not specific to human/Tbg infection. This particularly highlights the need to investigate the specificity of antigens used in these serologic tests for human *Trypanosoma* infections (14).

The RDT kit was designed to detect *T. b. gambiense* infections in humans (11), and has been deployed by the health authorities for HAT field surveillance/sampling in endemic countries. However, their inability to detect seropositive human samples and the detection of animal samples in this study location indicates significant shortcomings in the sensitivity and specificity of the RDT kit. This raises concerns about the reliability of surveillance studies conducted using this kit in Nigeria.

## Conclusion

The inability of the RDT kit to detect human seropositive samples while detecting positive animal samples in this study location highlights gaps in the sensitivity and specificity of HAT diagnostic tools used for field surveillance. This study emphasizes the need for more accurate field diagnostic tools for HAT, and for the validation of these tools in different endemic settings before deployment. Addressing these challenges will improve disease detection and reporting, as well as ensure appropriate policies and control measures towards the achievement of HAT elimination targets in 2030.

## Data Availability

All data produced in the present work are contained in the manuscript

## Abbreviations

CATT: Card Agglutination Test for Trypanosomiasis
ELISA: Enzyme Linked Immunosorbent Assay
HAT: Human African Trypanosomiasis
RDT: Rapid Diagnostic Test
VSG: Variant Surface Glycoprotein
ISG: Invariant Surface Glycoprotein

## Acknowledgments

We acknowledge the support of the African Academy of Sciences for the Vaccine Research Centre, University of Nigeria.

## Equal contribution

All authors contributed equally to this work

## Funding Statement

The authors did not receive any specific financial support for this work. However, CC is an APTI fellow of the African Academy of Sciences.

All authors’ current address: Vaccine Research Centre, University of Nigeria, Enugu, State, Nigeria.

## Notes

### Competing Interest Statement

The authors have declared no competing interest.

### Funding Statement

This study did not receive any funding

### Author Declarations

University of Nigeria Health Research Ethics Committee gave ethical approval for this work

## References

1. Papagni R, Novara R, Minardi ML, Frallonardo L, Panico GG, Pallara E, et al. Human African Trypanosomiasis (sleeping sickness): Current knowledge and future challenges. Frontiers in Tropical Diseases. 2023;4.

2. Ngutor KS, Idris LA, Oluseyi Oluyinka O. Silent Human Trypanosoma brucei gambiense Infections around the Old Gboko Sleeping Sickness Focus in Nigeria. J Parasitol Res [Internet]. 2016 [cited 2025 Apr 11];2016:2656121. Available from: https://pmc.ncbi.nlm.nih.gov/articles/PMC4752997/

3. Luintel A, Lowe P, Cooper A, MacLeod A, Büscher P, Brooks T, et al. Case of Nigeria-Acquired Human African Trypanosomiasis in the United Kingdom, 2016. Emerg Infect Dis. 2016;23(7):1225–1227.

4. Ozioko K, Okoye C, Obiezue R, Idika I, Awudu R, Ezewudo B, et al. Accelerating towards human African trypanosomiasis elimination: Issues and opportunities. J Vector Borne Dis [Internet]. 2020;57(2):105. Available from: 10.4103/0972-9062.310860

5. Camara O, Kaboré JW, Soumah A, Leno M, Bangoura MS, N’Diaye D, et al. Conducting active screening for human African trypanosomiasis with rapid diagnostic tests: The Guinean experience (2016–2021. PLoS Negl Trop Dis. 2024;18(2):11985.

6. Tablado Alonso S, Biéler S, Luz R, Verlé P, Büscher P, Hasker E. Retrospective clinical performance evaluation of the Abbott Bioline HAT 2.0, a rapid diagnostic test for human African trypanosomiasis based on recombinant antigens. Trop Med Int Health. 2025;30(2):135–142.

7. N’Djetchi MK, Camara O, Koffi M, Camara M, Kaba D, Kaboré J, et al. Specificity of serological screening tests and reference laboratory tests to diagnose gambiense human African trypanosomiasis: a prospective clinical performance study. Infect Dis Poverty. 2024;13, Articl.

8. Chappuis F, Loutan L, Simarro P, Lejon V, Büscher P. Options for field diagnosis of Human African trypanosomiasis. Clin Microbiol Rev. 2005;18(1):133–146.

9. Dukes P, Gibson W, Gashumba J, Hudson K, Bromidge T, Kaukus A, et al. Absence of the LiTat 1.3 (CATT antigen) gene in Trypanosoma brucei gambiense stocks from Cameroon. Acta Trop. 1992;51(2):123–134.

10. Meirvenne N, Magnus E, Büscher P. Evaluation of variant specific trypanolysis tests for serodiagnosis of human infections with Trypanosoma brucei gambiense. Acta Trop [Internet]. 1995;60(3):189–199. Available from: 10.1016/0001-706x(95)00127-z

11. C. L, P. B, P. L, S. B, S. B, J N. Performance of the SD BIOLINE® HAT rapid test in various diagnostic algorithms for gambiense human African trypanosomiasis in the Democratic Republic of the Congo. PLoS One. 2017;12(7):180555.

12. Eneh CI, Uwaezuoke SN, Okafor HU, Edelu B, Ogbuka S. Human African trypanosomiasis (sleeping sickness) in a Nigerian male adolescent and the treatment challenges: A case report. J Epidemiol Res. 2016;2(2).

13. Onah DN, Ebenebe OO. Isolation of a human serum-resistant Trypanosoma brucei from a naturally infected pig in the Nsukka area of Enugu State. Niger Vet J. 2004;24(1).

14. Ilboudo K, Hounyeme RE, Kabore J, Boulangé A, Gimonneau G, Salou E, et al. Experimental evidence that immune trypanolysis using the LiTat 1.3 and LiTat 1.5 variant antigen types is not specific to Trypanosoma brucei gambiense in pigs. Parasite. 2022;29.

15. Zongo K, Emmanuel RT. Advancing diagnosis and treatment for human African trypanosomiasis in Nigeria: challenges and future directions. Frontiers in Tropical Diseases. 2024;5(January):1–4.

16. Odebunmi EO, Ibeachu C, Chukwudi C.U. AO - Chukwudi CU, https://orcid.org/0000-0002-5118-5485 O. Prevalence of Human and Animal African Trypanosomiasis in Nigeria: a Scoping Review. medRxiv [Internet]. 2024; xAvailable from: https://www.medrxiv.org/

17. Jamonneau V, Camara O, Ilboudo H, Peylhard M, Koffi M, Sakande H, et al. Accuracy of individual rapid tests for serodiagnosis of gambiense sleeping sickness in West Africa. PLoS Negl Trop Dis [Internet]. 2015;9(2):3480. Available from: 10.1371/journal.pntd.0003480

18. Matovu E, Kitibwa A, Picado A, Biéler S, Bessell PR, Ndung’u JM. Serological tests for gambiense human African trypanosomiasis detect antibodies in cattle. Parasit Vectors. 2017;10(1).

